# Identification of modifiable plasma protein markers of cardiometabolic risk in children and adolescents with obesity

**DOI:** 10.1101/2025.02.25.25322850

**Authors:** Sara E. Stinson, Yun Huang, Roman Thielemann, Evelina Stankevic, Morten A.V. Lund, Louise A. Holm, Cilius E. Fonvig, Helene Bæk Juel, Dmitrii Borisevich, Maja Thiele, Aleksander Krag, Lars Ängquist, Thorkild I.A. Sørensen, Oluf Pedersen, Michael Christiansen, Jens-Christian Holm, Torben Hansen

## Abstract

Pediatric obesity is associated with multi-organ inflammation and an increased risk of cardiometabolic disease. To identify novel biomarkers of early-onset obesity-related cardiometabolic risk, we performed targeted proteomics, analyzing 149 proteins related to inflammation and cardiovascular disease in a cross-sectional sample of 2,377 children and adolescents with overweight/obesity and 1,647 with normal weight. In addition, we analyzed 64 circulating inflammation-related proteins in 184 children and adolescents, who participated in a 1-year family-based obesity management program. We identified a three plasma protein panel (CDCP1, FGF21 and HAOX1) that improved prediction of hepatic steatosis, compared with the predictive value of liver enzymes alone. In the prospective trial, we found that the non-pharmacological intervention induced a decline in circulating inflammatory cytokine levels, some of which were linked to improvements in cardiometabolic phenotypes. Our results may suggest that circulating proteomic signatures may mediate the associations between pediatric obesity and cardiometabolic risk.

## Main

The World Health Organization (WHO) has declared childhood obesity as one of the most serious public health challenges of the 21^st^ century, primarily due to the increasing global prevalence and the degree of associated cardiometabolic complications later in life, such as type 2 diabetes (T2D), metabolic dysfunction-associated steatotic liver disease (MASLD), and cardiovascular disease (CVD).^1^ The clustering of cardiometabolic risk factors—such as chronic low-grade inflammation, dysmetabolism and hypertension—can emerge early in life and serve as early predictors of future cardiometabolic comorbidities^2^. Hepatic steatosis is a particularly concerning complication of obesity in childhood, as it is associated with an increased risk of progression to cirrhosis in adulthood and an independent predictor of T2D risk^3^.

Pediatric obesity is highly heterogeneous, varying in clinical manifestation, progression, and treatment response. A key contributor to this heterogeneity is systemic inflammation^4^. Traditionally measured inflammatory markers, such as C-reactive protein, provide limited insights into the molecular pathways underpinning obesity-related metabolic dysfunction^4^. Advancements in targeted proteomics, such as the proximity extension assay (PEA), have enabled high-throughput quantification of low-abundance proteins, including cytokines, chemokines, and cardiovascular disease-related proteins^5^. This approach may unravel the mechanistic underpinnings of pediatric obesity, providing insights into the early stages of inflammatory and metabolic dysfunction. Despite this potential, this area remains relatively unexplored^6^. Large-scale studies on protein-disease associations have been conducted mainly in adult populations^7,8^, while studies in pediatric cohorts are more limited in sample size and differ in terms of methodology^9^ and scope^10^.

In this study, we applied PEA-based plasma proteomics alongside comprehensive phenotyping in children and adolescents from The HOLBAEK Study. Our aim was to investigate how circulating protein markers mediate the relationship between overweight/obesity and inflammation-related cardiometabolic risk. The cross-sectional analysis included 2,377 children and adolescents with overweight/obesity and 1,647 with normal weight^11,12^. Additionally, the study features a longitudinal intervention cohort of 184 children and adolescents with overweight/obesity who underwent the Holbaek obesity treatment—a family-centered, individualized, non-pharmacological approach involving regular follow-up visits at the Children’s Obesity Clinic, Holbaek Hospital, Denmark.

Our analysis identified distinct age-, sex-, puberty- and obesity-related plasma proteomic signatures linked to inflammation and metabolic disturbances from an early age. We uncovered protein marker patterns associated with hepatic steatosis, dyslipidemia, insulin resistance, hyperglycemia, hypertension, and related metabolic traits. Notably, reductions in BMI standard deviation scores (SDS) were associated with decreased circulating levels of inflammatory cytokines, highlighting the potential for interventions aimed at mitigating inflammation-related cardiometabolic dysfunction. This study not only advances our understanding of the proteomic landscape of pediatric obesity but may also lay the foundation for targeted prevention and individualized management of obesity-related comorbidities in children and adolescents.

## Results

### Study design

The study population was derived from The HOLBAEK Study^11,12^. It is composed of two cohorts: 1) an obesity clinic cohort, including children and adolescents with a BMI ≥90^th^ percentile (BMI SDS ≥1.28)^13^ who participated in a comprehensive, multidisciplinary non-pharmacological obesity management program at the Children’s Obesity Clinic, Holbæk Hospital; and 2) a population-based cohort, recruited from the school system across 11 municipalities in Zealand, Denmark.

Participant assessments included anthropometry, whole-body dual-energy X-ray absorptiometry (DXA) scanning^14^, proton magnetic resonance spectroscopy (^1^H-MRS)^15^ and blood biochemistry^12,16–25^, as previously described. PEA-based proteomics (Target 96 Cardiovascular II and Inflammation panels) were performed on 4,024 children and adolescents with baseline examinations and available blood sample (overweight/obesity, n = 2,377 and normal weight, n = 1,647), as well as a random subsample of the obesity clinic, including 184 participants who underwent obesity management and had a median follow-up period of 1.1 years (Target 96 Inflammation panel), as described previously^25^. An overview of the study design is presented in Fig 1.

**Fig 1.**
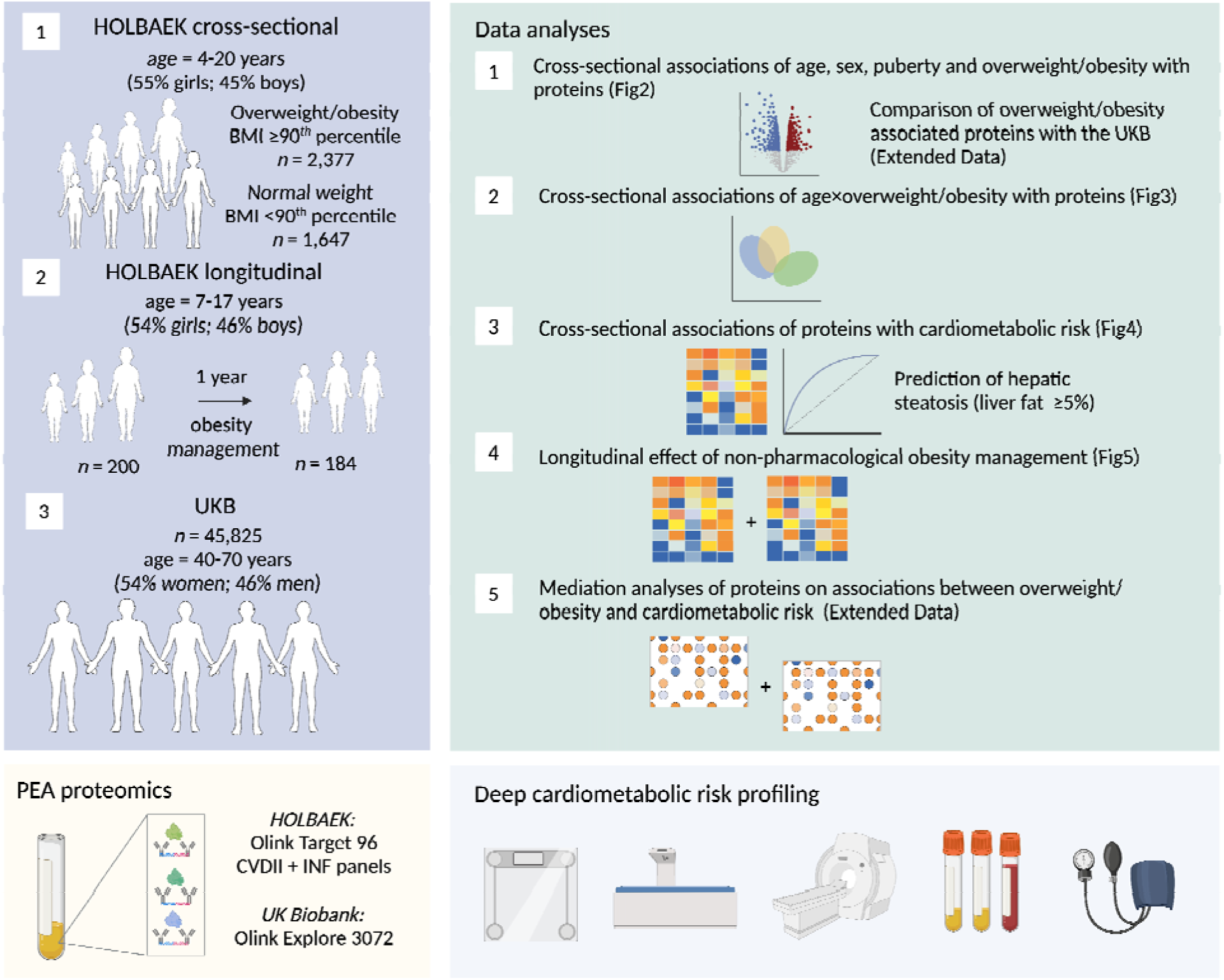
Study design. Proteomic profiles were measured using Olink Target 96 CVDII and INF panels in a cross-sectional study of 2,377 children and adolescents with overweight/obesity and 1,647 with normal weight. We assessed associations between circulating protein markers with age, sex and overweight/obesity, as well as with cardiometabolic risk profiles. We evaluated the predictive performance of circulating protein biomarkers for detecting hepatic steatosis. Additionally, the Olink INF panel was measured in a subset of 184 children and adolescents undergoing non-pharmacological obesity treatment. We examined changes in INF proteins in response to BMI SDS reduction and investigated longitudinal associations between protein marker changes and cardiometabolic risk profiles. BMI=body mass index; CVDII=cardiovascular II; INF=inflammation; PEA=proximity extension assay; SDS=standard deviation score.

### Study participant characteristics and cardiometabolic risk profiles

In the cross-sectional study, the group with overweight/obesity exhibited significantly higher cardiometabolic risk variables compared to the normal weight group (Extended Data Table 1). Specifically, the overweight/obesity group differed in terms of anthropometric measures, liver-related measures (liver fat % and routine liver function blood tests), lipid profiles, markers of general inflammation, glycemic indicators and blood pressure. Compared to their normal-weight counterparts, the overweight/obesity group had a higher prevalence of hepatic steatosis, defined as liver fat ≥5.0% (16.3% vs. 0%), elevated ALT levels (32.0% vs. 9.5%), dyslipidemia (21.7% vs. 5.9%), insulin resistance (44.5% vs. 6.8%), hyperglycemia (16.8% vs. 7.0%) and hypertension (14.9% vs. 5.6%), underscoring a high degree of cardiometabolic risk factor clustering.

### Associations of age, sex, and puberty with protein markers

Our analysis revealed significant associations between age and circulating protein levels, with 130 out of 149 proteins (87.2%) showing age-related differences independent of sex and BMI SDS (*P*<5% FDR; Fig 2a, Supplementary Table 1). Of these, 63 proteins were positively and 67 were negatively associated with age. Notably, circulating AGRP, LEP, and SPON2 levels increased with age. Conversely, proteins such as DNER, MMP12, and TNFRSF9 were more abundant at younger ages.

**Fig 2.**
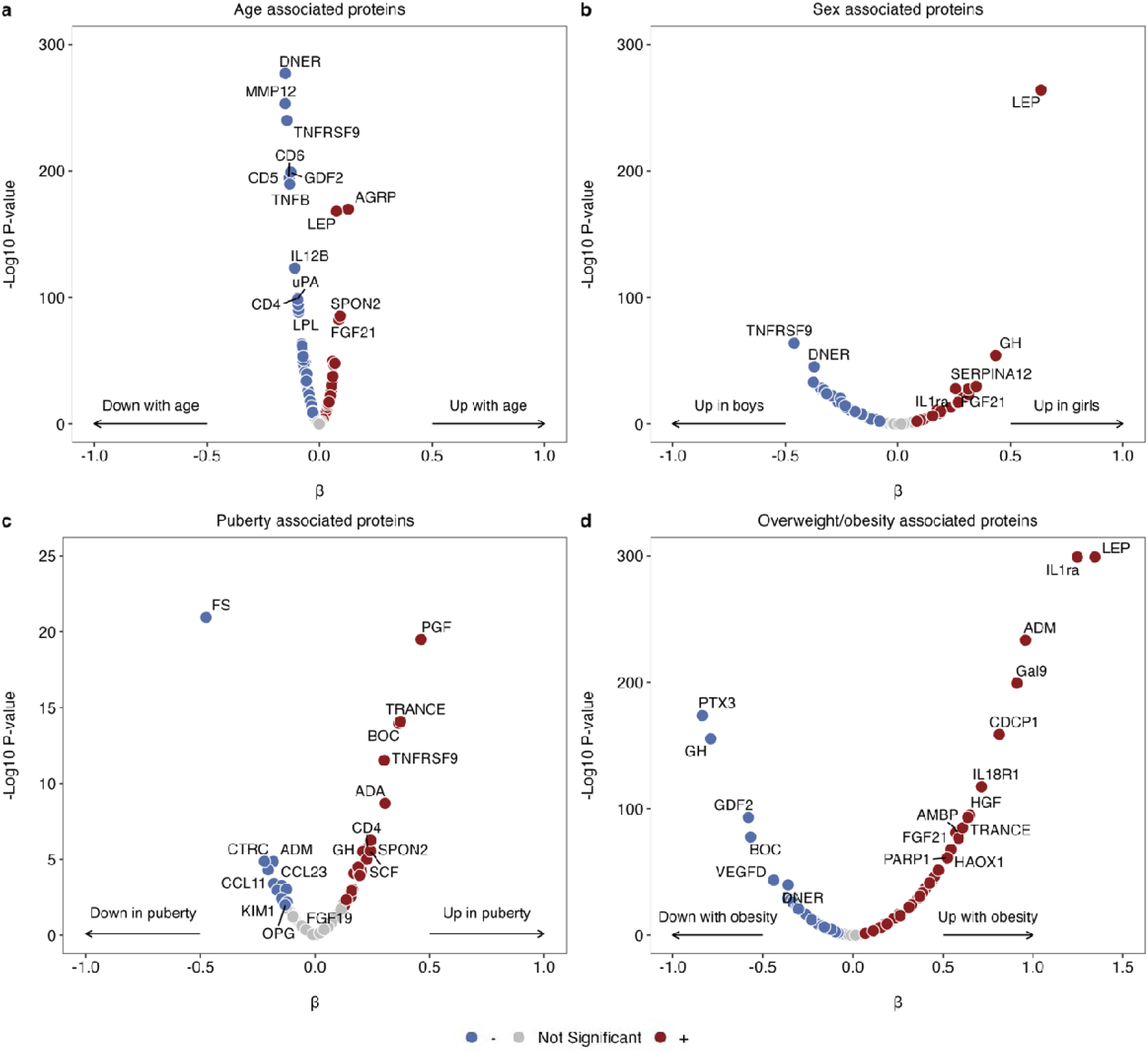
Plasma protein associations with age, sex and overweight/obesity. **a,** Significant associations of 130 out of 149 plasma proteins with age, adjusted for sex and BMI SDS. **b,** 83 out of 149 plasma proteins were associated with sex, adjusted for age and BMI SDS. **c,** 36 out of 149 plasma proteins were associated with puberty (Tanner stage), adjusted for age, sex, and BMI SDS. **d,** 122 out of 149 plasma proteins were associated with overweight/obesity compared to normal weight, adjusted for sex and age. The volcano plots show the estimated effect sizes from multiple linear regressions (n=2,377 with overweight/obesity and 1,647 with normal weight for age, sex, and BMI SDS as predictors; n=1,704 with overweight/obesity and 1,169 with normal weight for puberty as a predictor). Gray circles indicate non-significant (NS) associations, while red and blue represent positive and negative associations (P<5% FDR).

Sex-specific differences were observed for 83 out of 149 proteins (55.7%), independent of age and BMI SDS (Fig 2b, Supplementary Table 1). Of these, 31 proteins were significantly higher in girls and 52 were significantly higher in boys. Notably, girls exhibited higher levels of LEP, GH, and SERPINA12, while boys displayed elevated levels of TNFRSF9, PGF, and DNER.

Puberty-specific differences were observed for 36 out of 149 proteins (24.2%), independent of age, sex, and BMI SDS (Fig 2c, Supplementary Table 1). Of these, 22 proteins were significantly higher in puberty/post-puberty (Tanner stage = 2-5) and 14 were significantly higher in pre-puberty (Tanner stage = 1). Notably, puberty/post-puberty was associated with higher levels of PGF, TRANCE, and BOC, while pre-puberty was associated with higher levels of FS, CTRC, and ADM.

### Associations of overweight/obesity with protein markers

Overweight/obesity was associated with significant alterations in circulating protein levels, impacting 122 out of 149 proteins (81.9%), independent of age and sex (Fig 2d, Supplementary Table 1). Of these, 81 proteins were positively associated, and 41 were negatively associated with overweight/obesity compared to normal weight. Notably, LEP, IL1ra, ADM levels were higher with overweight/obesity and PTX3, GH and GDF2 levels were higher with normal weight.

To explore the age-specific nature of these associations, we compared protein markers linked to overweight/obesity in the pediatric HOLBAEK cohort with those observed in adults from the UK Biobank (UKB) (Supplementary Table 2). Although the direction of effects was largely consistent across age groups, key differences were noted in the magnitude of effects for certain proteins, including CDCP1, PTX3, IL1ra, GH, and TRANCE, which showed stronger estimated effect sizes in children and adolescents than in adults. Interestingly, some proteins displayed significant associations in the HOLBAEK pediatric cohort but not in the adult UKB. For example, OPG was inversely associated with overweight/obesity in the HOLBAEK cohort (β = - 0.25, *P* = 1.27E-15), but showed a non-significant association in UKB adults (β = 0.00, *P* = 0.61).

### Influence of overweight/obesity on age-related changes in protein markers

We examined age-related differences in circulating protein markers across weight groups using partial least squares-discriminant analysis (Fig 3a,b). Age groups were categorized into: Group 1 (girls <9 years, boys <10 years), Group 2 (girls 9–15 years, boys 10–16 years), and Group 3 (girls >15 years, boys >16 years)^26^. In individuals with normal weight, there was clear separation between age Groups 1 and 3, reflecting more distinct proteomic profiles across age groups. This separation was less pronounced in the group with overweight/obesity.

**Fig 3.**
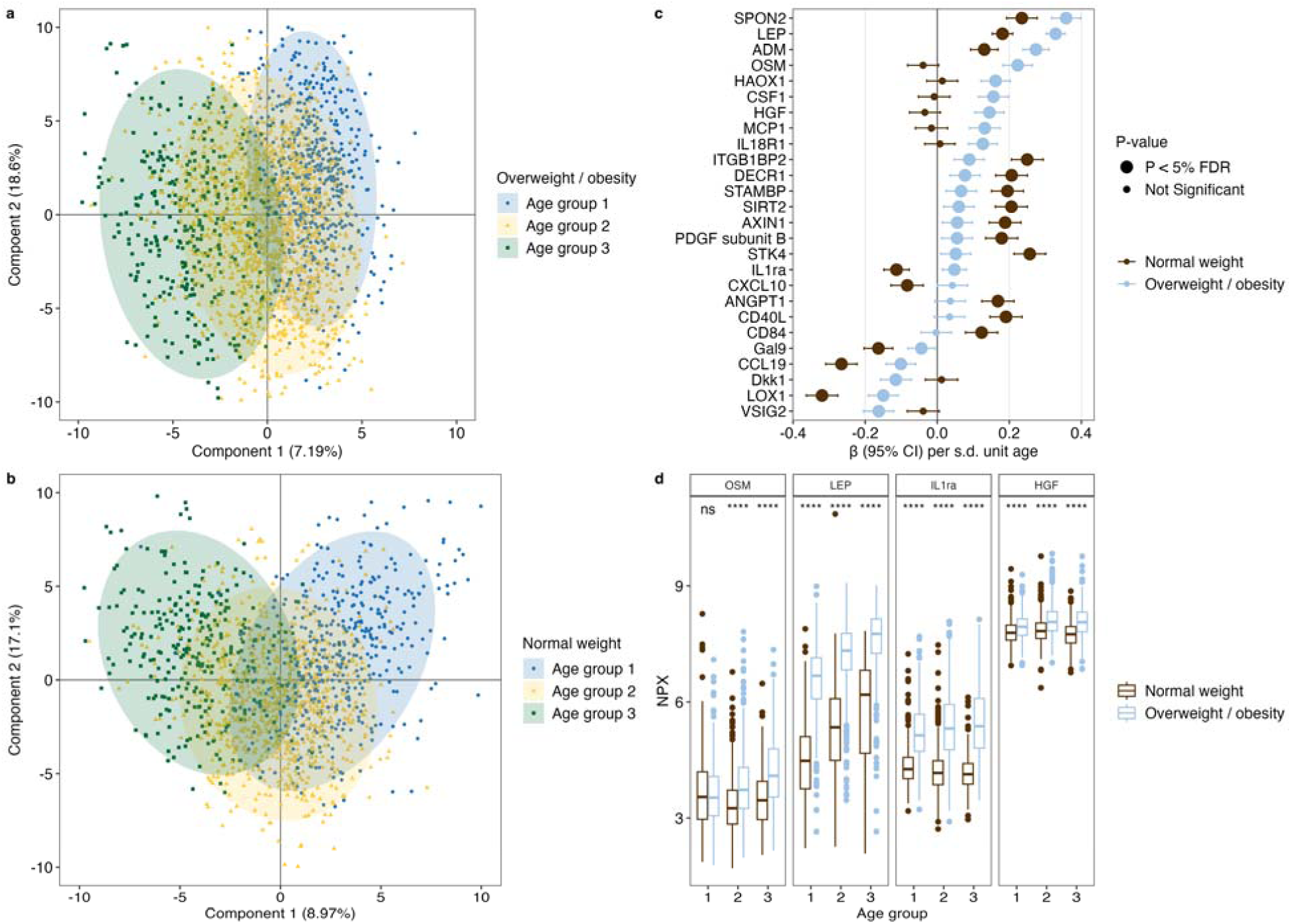
The interaction of overweight/obesity with age-associated circulating proteins. PLS-DA score plot showing plasma protein profiles across three age groups in children and adolescents with **a,** overweight/obesity and **b,** normal weight. **c,** Associations between age and top 26 proteins with significant age×overweight/obesity interactions (FDR P<1.0E-04). Linear regression models included an interaction term for age×overweight/obesity, adjusted for sex. β-coefficients with 95% CI are shown separately for the overweight/obesity (light blue; n = 2,377) and normal weight (brown; 1,647) groups. **d,** Box plots of the NPX levels for the four proteins with the largest age-related differences between overweight/obesity and normal weight, visualized across the three age groups. Three asterisks (***) indicate a significant difference between groups (P<0.001); ns, not significant. Sample sizes: n = 586, 1435, and 343 for age groups 1, 2, and 3 in the overweight/obesity group; n=486, 863, and 294 for age groups 1, 2 and 3 in the normal weight groups, respectively.

To further explore the influence of weight status on age-related changes in circulating protein markers, we included an interaction term (age×[overweight/obesity vs. normal weight]) in regression models for each protein, adjusting for sex. We identified significant interactions (*P <5*% FDR) for 90 out of 149 proteins (60.4%), indicating that the relationship between age and protein levels also differs by weight status (Supplementary Table 3). The top 26 interactions (FDR *P*<1.0E-04) are illustrated in Fig 3c. Notably, several proteins showed amplified age-related increases in the overweight/obesity group compared to the normal weight group. For example, OSM exhibited a positive age-related trend in the overweight/obesity group (β = 0.22), yet a slight negative trend in the normal weight group (β = -0.04, *P*_Interaction_ = 6.23E-18), same with HGF (β_overweight/obesity_= 0.14 vs. β_normal weight_ = -0.03, *P*_Interaction_ = 2.32E-09). Similarly, LEP (β_overweight/obesity_ = 0.33 vs. β_normal weight_ = 0.18, *P*_Interaction_ = 1.37E-13) and IL1ra (β_overweight/obesity_ = 0.05 vs. β_normal weight_ = -0.11, *P*_Interaction_ = 7.96E-11) showed stronger age-related increases.

To pinpoint when these age×overweight/obesity interactions emerge, we examined differences in these protein markers across the three age groups according to overweight/obesity or normal weight (Fig 3d). Most markers were higher across all three age groups. Notably, OSM displayed a unique pattern, with significant differences only between age Groups 2-3, suggesting a potential age-related shift in protein expression during puberty, which may be influenced by weight status.

### Protein marker associations with cardiometabolic risk profiles

Of the 149 proteins analyzed, 128 proteins were significantly associated with at least one dichotomized cardiometabolic risk feature (*P*<5% FDR), independently of age, sex and BMI SDS (Supplementary Table 4). Specifically, 118 proteins were linked to a higher prevalence of cardiometabolic conditions, including hepatic steatosis (liver fat ≥5%), elevated ALT, dyslipidemia, insulin resistance, hyperglycemia and hypertension (Fig 4a, Extended Data Fig 1).

**Fig 4.**
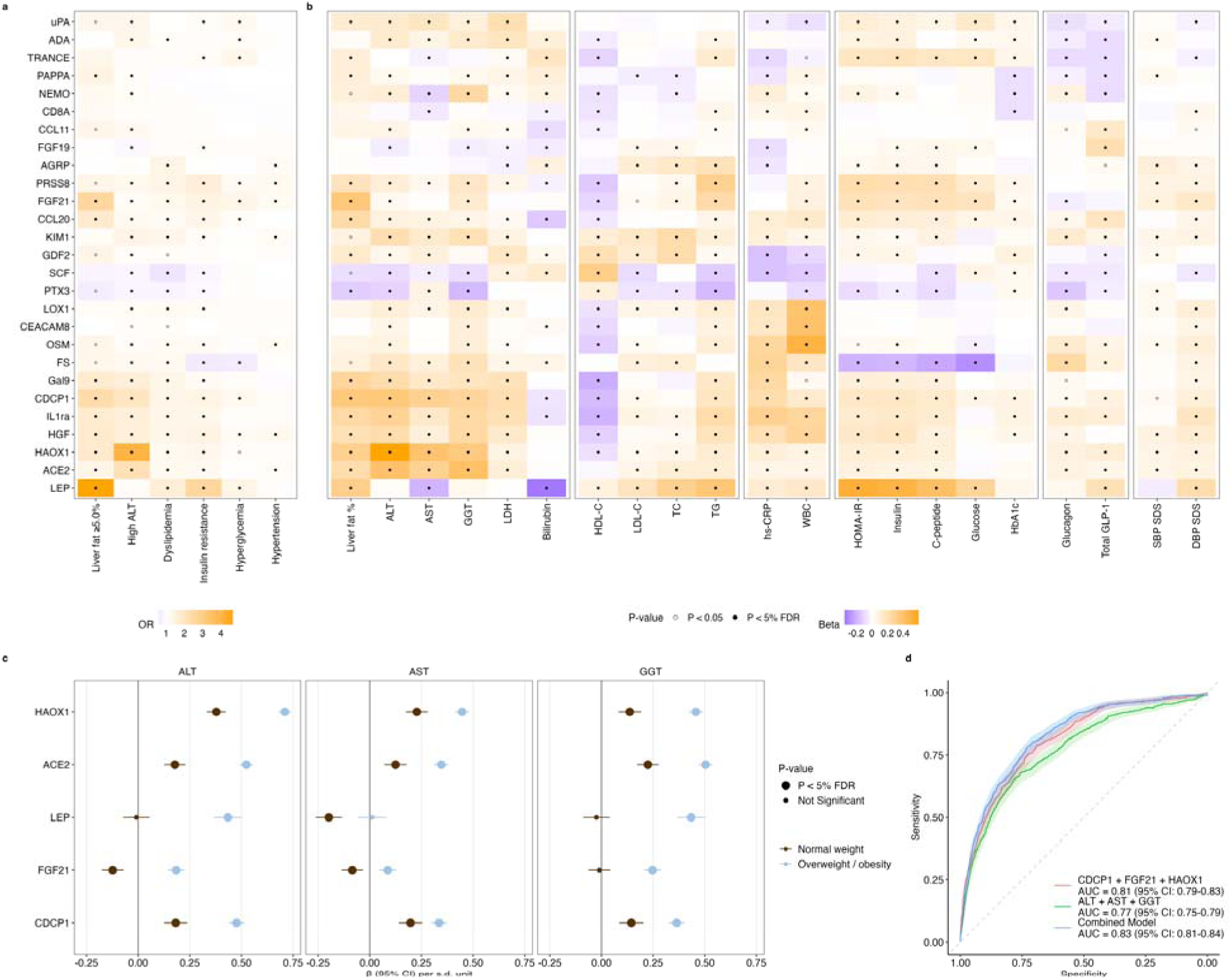
Associations of plasma proteins with cardiometabolic risk profiles. **a,** Top 27 plasma proteins associated with cardiometabolic risk features (P <5% FDR), identified through logistic regression analysis, adjusted for age, sex, and BMI SDS. **b,** Associations of top proteins with cardiometabolic traits, tested with linear regression, adjusted for age, sex, and BMI SDS. **c,** Associations between five proteins and liver enzymes with significant overweight/obesity interactions (P<5% FDR). Linear regression models included an interaction term for overweight/obesity vs. normal weight, adjusted for age and sex. β-coefficients with 95% CI are shown separately for the overweight/obesity (light blue; n=2356) and normal weight (brown; n=1619) groups. **d,** Discriminant accuracy of three proteins and liver enzymes for diagnosing hepatic steatosis (liver fat ≥5.0%). Analysis was performed on 425 participants with controls downsampled to match the 76 cases of hepatic steatosis. Each ROC curve is accompanied by its corresponding 95% CI (shaded area), and the mean AUC values with their respective 95% CIs are provided for each curve.

For continuous phenotypes, all 149 proteins had at least one significant association (*P*<5% FDR) with a cardiometabolic trait, independently of age, sex, and BMI SDS (Fig 4b; Supplementary Table 5; Extended Data Fig 1). For *liver-related traits*, examples of top significant positive predictors of ALT levels include HAOX1, ACE2, and CDCP1. For *lipid-related traits*, PRSS8, FGF21 and AMBP emerged as the strongest positive predictors for plasma triglyceride levels. For *glycemic-related traits*, LEP and PRSS8 emerged as the strongest positive predictors of insulin, on the other hand FS was the strongest negative predictor. In general, FS was inversely associated with glycemic traits, yet positively associated with most other cardiometabolic risk factors. For *inflammation-related traits*, OSM, CEACAM8, and LOX1 were the most notable positive predictors of WBC. For *proglucagon hormones*, we observed FGF19, CCL20 and CCL11 among strongest positive predictors for total glucagon-like peptide-1 (GLP-1). For *blood pressure-related traits*, FGF21, IL1ra, and LEP were identified as the strongest positive predictors of diastolic blood pressure.

### Sex-specific associations of protein markers with cardiometabolic traits

We investigated sex differences in the associations between circulating proteins and cardiometabolic traits, revealing significant sex interactions (*P<5*% FDR) for 64 out of 149 proteins (43.0%) across 13 of 21 cardiometabolic traits, independently of age and BMI SDS (Supplementary Table 6). Notably, several proteins exhibited opposite directions of effect between girls and boys. For example, TNFRSF9 was positively associated with ALT levels in girls (β = 0.14) but negatively in boys (β = -0.15; *P_Interaction_* = 8.61E-19). Similarly, FGF21 (β*_girls_* = -0.05 vs. β*_boys_* = 0.15, *P_Interaction_* = 8.35E-11), CD4 (β*_girls_* = 0.14 vs. β*_boys_* = -0.07, *P_Interaction_* = 2.13E-10) and TWEAK (β*_girls_* = 0.04 vs. β*_boys_* = - 0.15, *P_Interaction_* = 1.24E-10) showed sex-dependent associations with ALT levels.

### Influences of overweight/obesity in protein marker-cardiometabolic associations

To investigate the role of overweight/obesity in modulating the associations between protein levels and cardiometabolic traits, we assess interaction effects (protein×[overweight/obesity vs. normal weight]), independent of age and sex (Supplementary Table 7). Significant interactions were observed for 127 out of 149 proteins (85.2%) across 19 out of 21 cardiometabolic traits (*P*<5% FDR). For *liver-related traits*, the strongest interactions with overweight/obesity on ALT levels were observed for HAOX1, ACE2, LEP, FGF21 and CDCP1. Notably, FGF21 showed opposite direction of effects between weight groups (**Fig 4c**). For *lipid-related traits*, LEP, ADM, and PTX3 had the most significant interactions with overweight/obesity for triglycerides. For *glycemic-related traits*, the strongest interactions with overweight/obesity were observed for LEP, ADM, and SCF with fasting serum insulin levels. For *inflammation-related traits*, LEP, FGF21 and IL1ra had the most significant interactions with overweight/obesity for WBC levels. For *proglucagon hormones*, LEP, FGF21, and HGF had the most significant interactions with overweight/obesity for fasting glucagon. For *blood pressure-related traits*, LEP, ADM and DNER emerged as the strongest interaction with overweight/obesity for diastolic blood pressure.

### Circulating protein marker panel for prediction of hepatic steatosis

To improve non-invasive detection of hepatic steatosis, defined as liver fat exceeding 5%, we utilized elastic net regularization for feature selection, identifying a three-protein panel consisting of CDCP1, FGF21, and HAOX1. This panel achieved a mean cross-validated area under the receiver operating characteristic curve (ROC-AUC) of 0.81 (95% CI: 0.79-0.83) using five-fold cross-validation repeated ten times (Fig 4d). Notably, combining this protein panel with traditional liver enzymes (ALT, AST, and GGT) significantly increased the AUC from 0.77 (95% CI: 0.75–0.79) to 0.83 (95% CI: 0.81– 0.84), as confirmed by DeLong’s test (*P*<0.05). These findings underscore the added predictive value of circulating protein biomarkers in assessing hepatic steatosis risk.

### Mediation effect of protein markers on cardiometabolic traits

To investigate the mechanistic role of circulating protein markers in the relationship between overweight/obesity and cardiometabolic traits, we performed mediation analysis, adjusted for age and sex. Out of 149 proteins analyzed, 122 significantly mediated the effect of overweight/obesity on 21 cardiometabolic traits (*P*<5% FDR), with a median proportion of 2%. Notably, 38 proteins exhibited substantial mediation effects, exceeding more than 20% of the total effect. Key protein mediators include LEP, IL1ra, IL18R1, CDCP1, HAOX1, and HGF which influenced a broad range of cardiometabolic traits, highlighted from hierarchical clustering (Extended Data Fig 2 and Supplementary Table 8).

### Protein marker changes in response to individualized pediatric obesity treatment

We evaluated protein marker changes in response to personalized obesity treatment among 184 children and adolescents (84 boys) with overweight/obesity, with a median age of 11.6 years (IQR: 9.9–13.5). Participants with complete data on BMI SDS and blood samples at baseline and follow-up were included, with a median follow-up duration of 1.1 years (IQR: 1.0–1.2). Of these, 151 showed a reduction in BMI SDS, while 33 maintained or increased their BMI SDS. The median decrease in BMI SDS was −0.39 (IQR: −0.76 to −0.07), accompanied by significant improvements in body fat percentage, lipid levels, glucose, HbA1c and blood pressure (all *P*<0.05; Extended Data Table 2), as previously reported^25^.

We next explored the relationship between BMI SDS reduction and changes in inflammatory (INF) protein levels, independently of age, sex and treatment duration. A total of 14 out of 64 INF proteins were significantly associated with BMI SDS reduction (*P*<5% FDR). Notably, IL18R1, IL18, FGF21, IL10RB, and CDCP1 showed the largest decrease following BMI SDS reduction (Fig 5a, Supplementary Table 9).

**Fig 5.**
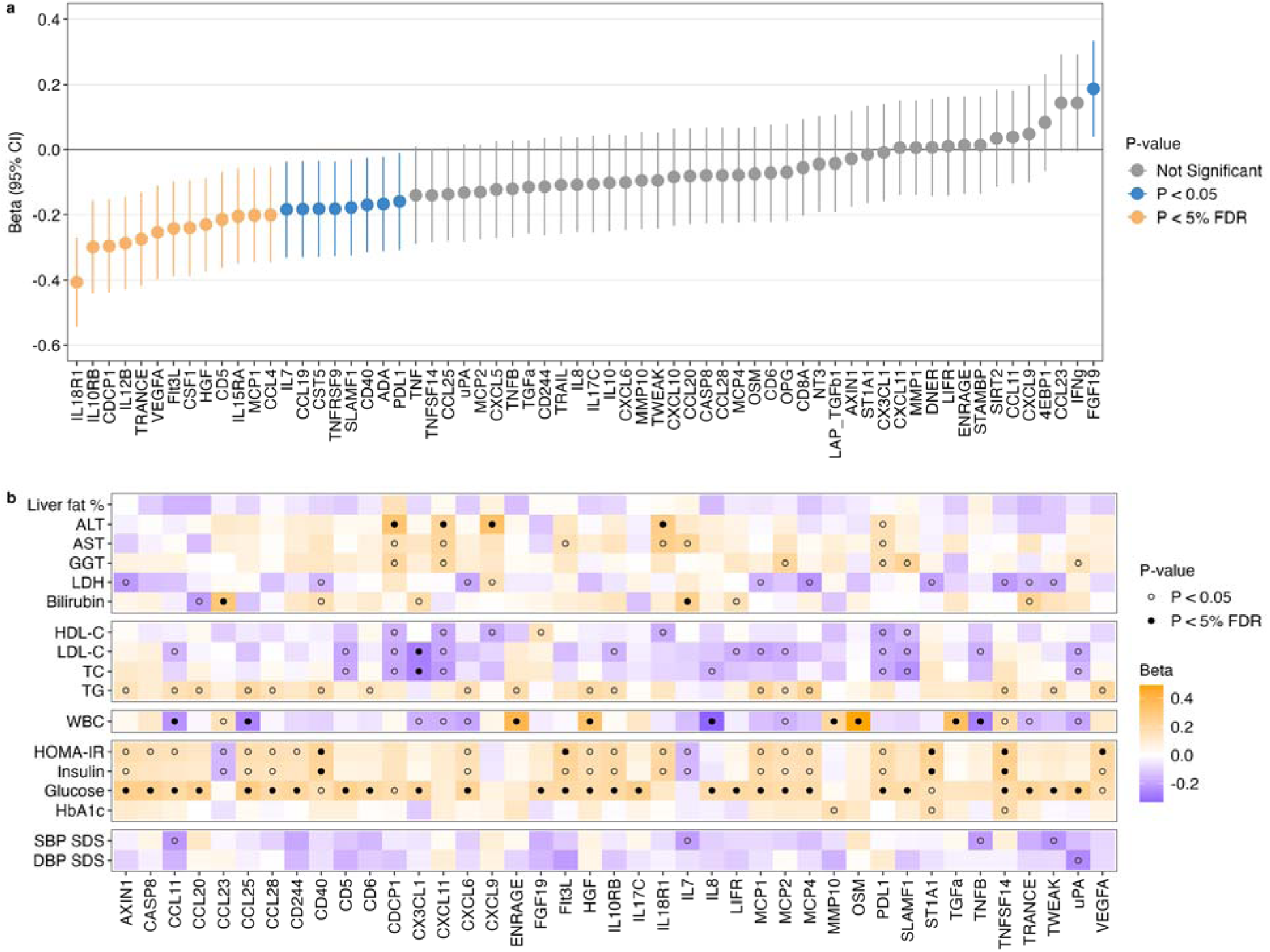
Non-pharmacological obesity treatment and changes in protein marker levels and cardiometabolic risk profiles. **a**, BMI SDS reduction was associated with changes in 14 INF proteins (P<5% FDR), assessed using linear regression, adjusted for age, sex and treatment duration. The β-coefficients with 95% CIs are shown for each protein. **b**, Changes in INF proteins were associated with changes in cardiometabolic traits, tested by linear regression, adjusted for age, sex, treatment duration, baseline BMI SDS and changes BMI SDS. BMI SDS reduction was calculated as the difference between baseline BMI SDS and follow-up values. Changes in INF proteins and cardiometabolic traits were calculated as the difference between follow-up and baseline values (n=184, P<5% FDR).

Longitudinal analyses revealed 62 significant associations between 47 out of 64 INF proteins and eight out of 17 cardiometabolic risk factors (*P*<5% FDR), independently of age, sex, treatment duration, baseline BMI SDS and changes BMI SDS (Fig 5b, Supplementary Table 10). *For liver-related traits*, decreases in IL18R1, CDCP1, CXCL11 and CXCL9 levels were linked to reductions in ALT levels. For *lipid-related traits*, changes in CX3CL1 levels were negatively associated with changes in LDL-C and TC. Numerous INF protein changes were associated with changes in *glycemic-related traits*. Decreases in TNFSF14, ST1A1, CD40, Flt3L and VEGFA levels were linked to reductions in HOMA-IR levels. For *inflammation*, decreases in HGF, OSM, TGFa, ENRAGE and MMP10 were associated with reductions in WBC levels. No INF proteins were significantly associated with changes in *blood-pressure related traits*, after multiple testing corrections.

### Mediation effect of changes in protein markers on cardiometabolic traits

We investigated whether changes in protein markers mediate the relationship between BMI SDS reduction and changes in cardiometabolic traits, independently of age, sex, and treatment duration (Extended Data Fig 3, Supplementary Table 11). Changes in 18 proteins significantly mediated the association between BMI SDS reduction and changes in 12 out of 17 cardiometabolic risk factors (*P*<5% FDR). Notably, IL18R1 and CDCP1 were key positive mediators in the relationship between BMI SDS reduction and improvements in *liver-* and *glycemic-related traits*. Interestingly, CDCP1 exhibited inverse mediation effects on lipid levels, suggesting a complex role in lipid metabolism.

## Discussion

Our study highlights the profound impact of overweight and obesity on altered expression of circulating protein markers in children and adolescents, revealing strong associations with cardiometabolic risk profiles. These findings contribute to a growing body of evidence that pediatric obesity increases systemic inflammation and disrupts metabolic pathways, contributing to increased risk of developing cardiometabolic comorbidities. We observed that nearly half of children and adolescents with overweight or obesity exhibited insulin resistance (45%), with notable proportions also exhibiting dyslipidemia (22%), hyperglycemia (17%), hepatic steatosis (16%), and hypertension (15%). These results emphasize the urgent need for early interventions aimed at reducing cardiometabolic risk to mitigate the long-term consequences of pediatric obesity.

Prior studies in pediatric populations are limited, yet with advances in proteomics technology and large-scale biobanks, we now have the power to identify early markers of cardiometabolic dysfunction. Recent work from The HOLBAEK Study has begun to map proteomic changes related to age, sex, and BMI SDS in youth using mass-spectrometry (MS)-based approaches, quantifying approximately 1,200 plasma proteins in 3,147 children and adolescents^9^. The present study further advances this field, utilizing PEA technology to capture lower-abundant proteins, such as inflammatory markers, which remain challenging to quantify using MS methods^5^. Similar to the present study, age-related shifts in bone markers, such as OSM differed with obesity, similar findings highlighted by previous MS study^9^, including declines in aggrecan levels — a key protein in bone development^27^.

Additional pediatric biobanks include the ALSPAC cohort, of which 92 proteins have been measured in non-fasted plasma samples using the Olink Target Inflammation panel, in 3,005 9-year-olds, although the results of this study have not yet been published^10^. Our findings also align with studies performed in adults, including those from the BAMSE cohort (2,074 adults; mean age of 22.5 years)^28^ and the UK Biobank (46,825 adults; mean age of 56.5 years)^8^. In the present study, we demonstrated consistent directions of effects of adiposity associated with circulating plasma proteins across childhood (HOLBAEK) and adulthood (UKB). Strikingly, some proteins exhibited age-specific associations, present only in childhood and adolescence, particularly those related to factors such as bone metabolism. For example, OPG —a known marker of bone strength^29^— was inversely associated with overweight/obesity in the HOLBAEK cohort, but was non-significant in UKB adults, highlighting certain proteins that may have age-specific effects during growth and development.

In the present study, we identified key protein signatures associated with cardiometabolic risk patterns. For *liver-related* phenotypes, we observed HAOX1, ACE2, and CDCP1 to be among the strongest positive predictors of plasma ALT levels, proteins previously associated with adult MASLD^30–32^. For *lipid-related* traits, proteins such as PRSS8 were significantly associated with triglyceride levels. Notably, variants in the *PRSS8* gene have previously been implicated in lipid metabolism through a genome-wide association study^33^. We observed a divergent pattern across cardiometabolic traits for FS (follistatin), which was inversely associated with *glycemic traits*, yet positively associated with most other cardiometabolic risk factors. Folkersen *et al.*, also highlighted an inverse causal effect of FS levels on fasting glucose and insulin, through Mendelian randomization analysis^34^. For traditional *inflammation-related* phenotypes, we observed significant associations between hs-CRP and WBC with markers such as LOX1, previously considered a novel biomarker for ischemic stroke and a potential drug target^35^. Prior work from our group found LOX1 to be associated with inflammation in pediatric obesity^23^. For *proglucagon hormones*, we observed strong interactions between overweight/obesity and proteins including LEP, FGF21, and HGF on fasting glucagon levels, which may highlight the potential role of the liver-alpha-cell axis in obesity-related metabolic dysregulation^36^. We also observed significant links between several inflammatory proteins, such as FGF19, CCL20 and CCL11 and fasting GLP-1 levels, which may fit with the theory that GLP-1 has more diverse functions, beyond simply stimulating insulin secretion, including modulating response to inflammation^37^. For *blood pressure-related* traits, we observed certain proteins, including FGF21 and IL1ra significantly associated with diastolic blood pressure (DBP) SDS. FGF21 levels are associated with hypertension in adults^38^ and IL1ra antagonists are in clinical trials to reduce blood pressure^39^.

A key finding of our study was the identification of a three-protein panel— CDCP1, FGF21, and HAOX1—that outperformed traditional liver enzymes in predicting hepatic steatosis (defined by ≥5.0% liver fat), with an area under the curve (AUC) of 81% compared to 77% for liver enzymes alone. The combination of the protein panel with liver enzymes further improved diagnostic accuracy to 83%. CDCP1 has emerged as a particularly promising biomarker, with previous studies reporting elevated levels in adults with metabolic dysfunction-associated steatohepatitis (MASH)^30^ and in pediatric obesity^40^. Its association with obesity and cardiometabolic risk factor clustering, alongside its decrease in levels following BMI SDS reduction during the intervention, highlights its potential role in monitoring treatment response. Similarly, FGF21 and HAOX1 were strongly linked to obesity and liver disease markers in the present study, underscoring their relevance as non-invasive biomarkers for metabolic health in a pediatric population. Moreover, previous work from our group has identified a panel of lipids predictive of hepatic steatosis^25^ to a similar extent as the current study, suggesting that subsequent combination of data across omics layers, may lead to improved predictive performance. The clinical utility of proteomics-based scores is further highlighted by prior studies performed in adults^41,42^. Future studies should validate this three-protein panel in external cohorts, ideally with biopsy-confirmed MASLD to confirm its clinical utility.

While proteomic profiling offers valuable insights into the biological underpinnings of pediatric obesity, heterogeneity in treatment response remains a critical challenge^1^. Despite successful weight loss in over 80% of participants, approximately 20% were non-responders, highlighting the complexity of obesity management. Our data revealed significant decreases in 14 inflammatory proteins with BMI SDS reduction, which correlated with improvements in cardiometabolic risk factors during the intervention. Notably, reductions in IL18, IL18R1, FGF21, and CDCP1 were linked to improved glycemic traits and liver enzyme profiles, suggesting potential pathways for targeted therapeutic strategies. However, the persistence of elevated levels of certain proteins indicates that lifestyle interventions alone may be insufficient for all patients. Pharmacological approaches, such as GLP-1 receptor agonists, could complement personalized obesity management by targeting specific inflammatory pathways^43^.

This study has limitations that warrant consideration. Our cohort was predominantly of European ancestry, limiting generalizability to more diverse populations. Additionally, the targeted proteomics approach restricted the analysis to proteins captured by the PEA panels, potentially overlooking other relevant biomarkers. We did not account for genetic predispositions or lifestyle factors, such as diet and physical activity, which likely influence circulating protein levels. The relatively small number of hepatic steatosis cases, warrant cautious interpretation of the predictive model’s performance. Furthermore, the absence of a control group in the longitudinal analysis limits causal inferences regarding the impact of the obesity management intervention. Lastly, we have compared proteomics data between the HOLBAEK Study and UKB, which used two slightly differing methodologies, Olink Explore 3072 vs. Target 96 platforms. Additional cohort specific differences may exist, including intake of medications, which is more common in adults and may affect circulating protein expression. Nevertheless, our study’s strengths lie in the inclusion of deeply phenotyped cohorts of children and adolescents from both clinical and population-based settings, as well as the integration of both cross-sectional and longitudinal intervention data, with comparison to an adult cohort. This comprehensive approach enhances the robustness and clinical relevance of our findings.

In conclusion, our study highlights the potential of proteomic profiling to identify novel circulating biomarkers implicated in the pathophysiology of pediatric obesity and its associated cardiometabolic comorbidities. The three-protein panel for hepatic steatosis offers a promising non-invasive diagnostic tool, paving the way for earlier detection and possible intervention. Importantly, personalized obesity management can modulate circulating protein levels and cardiometabolic risk, though a subset of patients may require adjunctive pharmacotherapy. Future research should focus on validating these findings in diverse populations, exploring integration across various omics layers, and developing precision medicine strategies tailored to pediatric obesity phenotypes.

### Online Methods Ethics

According to the Declaration of Helsinki, written informed consent was obtained from all participants. An informed oral assent was given by the participant if the participant was younger than 18 and then the parents gave informed written consent. The study was approved by the Ethics Committee of Region Zealand, Denmark (SJ-104) and by the Danish Data Protection Agency (REG-043-2013). The HOLBAEK study is registered at ClinicalTrials.gov (NCT00928473).

### Study population

Both cohorts from the HOLBAEK study were enrolled between January 2009 and April 2019. Among 4,133 children and adolescents with available plasma proteomics in the cross-sectional study, participants were excluded based on diagnosed type 1 diabetes mellitus or T2D (*n*=7); intake of medications including statins, insulin, metformin and liraglutide (*n*=8); meeting T2D criteria based on blood sample (fasting plasma glucose ≥7.0mmol l^−1^ and/or HbA1c ≥48mmol mol^−1^) (*n*=8); the interval between visit and blood sample collection >90 days (*n*=30); underweight (BMI <5th percentile [BMI SDS <−1.64]) (*n=*53). As a result, 4024 participants were included in the cross-sectional analysis.

Children and adolescents with overweight or obesity (BMI SDS ≥1.28) enrolled in the multidisciplinary, family-based and individual-centered obesity clinic cohort received comprehensive management using an evidence-based treatment protocol which comprises a range of recommendations on nutrition, including meal exercises, picky eating, exercise, inactivity, border setting, promoting growth, development and improved physical, mental and social thriving, as previously described^11^. The intervention study included 200 children and adolescents with overweight or obesity, who were followed for a median of 1.1 years (IQR 1.0–1.2) and 184 had available blood samples at follow-up. Their proteomic profiles were available at both baseline and follow-up.

### Anthropometric measurements

In the obesity clinic cohort, anthropometrics were obtained at clinical examinations, whereas the population-based group was assessed in a mobile laboratory by trained medical professionals^22^. Weight, height, waist, and WHR were measured. BMI SDS was calculated according to a Danish reference^13^, waist SDS and waist-hip SDS was calculated according to age- and sex-specific reference values^44^. For systolic blood pressure (SBP) and DBP, mean values for the last two measurements of BP were calculated and converted to BP SDS, based on age-, sex-, and height-specific reference values from the American Academy of Pediatrics^45^.

### Puberty stage

Tanner stage^46,47^ was evaluated by trained medical professionals in the obesity clinic cohort and self-reported using a questionnaire with picture pattern recognition in the population-based cohort. Puberty stage was then classified as either Pre-pubertal = Tanner stage 1 or Pubertal = Tanner stage 2–5.

### Proteomics

PEA was performed using the Target 96 Cardiovascular II (CVDII) and Target 96 Inflammation (INF) panels at baseline from Olink Proteomics on EDTA plasma, as previously described^23–25^. PEA technology uses nucleic acid labeling of antibodies in combination with qPCR, producing normalized protein expression values as an arbitrary unit on a log2 scale. Overall, 85 markers from CVDII and 64 markers from INF for a total of 149 proteins were included as >80% of individuals were above the limit of detection (LOD; Extended Data Table 3). Each Protein was inverse-rank normalized, including NPX data below the LOD, prior to analyses and association testing.

### DXA examination

Whole-body DXA scans were performed and total body fat percentage was quantified in the overweight/obesity (*n*=1,330), normal weight (*n*=113) groups and 124 children and adolescents with overweight or obesity who received the obesity management, using a GE Lunar Prodigy (DF+10031, GE Healthcare) until October 2009 and thereafter using a GE Lunar iDXA (ME+200179, GE Healthcare)^14^.

### 1H-MRS examination

Liver fat content was quantified in the overweight/obesity (*n*=479) and normal weight (*n*=34) groups, as well as 96 children and adolescents with overweight/obesity who received obesity management, using a 3T Achieva MR imaging system (Philips Medical Systems), as previously described^15^. Data processing was performed by a trained magnetic resonance physicist.

### Biochemical analyses

Venous blood samples were collected after an overnight fast. Fasting biochemical measurements include in plasma: alanine transaminase (ALT), aspartate transaminase (AST), γ-glutamyl transferase (GGT), lactate dehydrogenase (LDH) and bilirubin^16^, high-density lipoprotein cholesterol (HDL-C), low-density lipoprotein cholesterol (LDL-C), total cholesterol (TC), triglycerides (TG)^20^, glucose^17^, glucagon^19^ and GLP-1^18^, in serum: insulin, C-peptide^17^, high-sensitivity C-reactive protein (hs-CRP)^22^, and in whole blood: hemoglobin A1c (HbA1c)^17^ and white blood cell count (WBC)^22^, as previously described.

### Defining cardiometabolic risk features

Hepatic steatosis was defined using the histological cutoff of ≥5.0% liver fat in adults^48^. We also defined high ALT (above 24.5 U/L in girls and above 31.5U/L in boys), which was found to be the optimal cutoff for diagnosing hepatic steatosis by our group^16^. Hyperglycemia was defined as fasting plasma glucose ≥5.6 to 6.9mmol/L and/or HbA1c ≥39 to 47mmol/mol, according to the American Diabetes Association guidelines for prediabetes^49^. Insulin resistance was defined based on HOMA-IR value above the 90th percentile of previously published age- and sex-specific population-based reference values from our group^17^. Homeostasis model assessment of insulin resistance (HOMA-IR) was calculated as (insulin mU/L x glucose mM) / 22.5. Dyslipidemia was defined as values above the 95th percentile according to pediatric guidelines, corresponding to: TC ≥200mg/dL (5.2mM), LDL-C ≥130mg/dL (3.4mM), TG ≥100mg/dL (1.1mM) for 0-9 years or ≥130 mg/dL (1.5mM) for 10-19 years, or HDL-C <40mg/dL (1.0mM)^50^. Hypertension was defined as a SBP and/or DBP above the 95th percentile for age, sex, and height^45^.

### Statistical analyses

Statistical analyses were performed using R software version 4.4.1^51^. Descriptive summary data are expressed as median (IQR) for continuous variables or frequencies and percentages for categorical variables. The Wilcoxon rank-sum test (for continuous variables) and the chi-squared test (for categorical variables) were used to test for differences in characteristics between two groups.

Multiple linear regressions were used to examine the association of age, sex, puberty or overweight/obesity versus normal weight with each protein individually. PLS-DA was performed to examine how protein profiles differ between the three age groups, separately for overweight/obesity versus normal weight groups using *ropls* v.1.36.0 R package^52^. Ten-fold cross-validation and 300 permutations were used. The first two component scores were plotted in a score plot, where each point represents an individual.

The effect of obesity on the association between continuous age and individual circulating protein markers were tested by a corresponding interaction model including an interaction term (age×overweight/obesity vs. normal weight) adjusting for sex.

Cardiometabolic traits were log-transformed except for BMI SDS, SBP SDS and DBP SDS. The associations of protein biomarkers with cardiometabolic risk features and cardiometabolic traits were examined using multiple logistic and linear regressions adjusted for age, sex and BMI SDS when pooling the overweight/obesity and normal weight groups. The respective interactions between sex and protein biomarkers (protein×sex) and obesity (protein×overweight/obesity vs. normal weight) were examined by linear regression for cardiometabolic traits, adjusted for age and either BMI SDS or sex. The reported estimates (β or OR) are based on a 1-s.d. unit increase in independent variables. Multiple testing correction was performed based on FDR at 5% to be considered statistically significant. In all figures, only those proteins with at least one outcome association reaching FDR significance were included.

Changes in protein profiles before and after obesity management were assessed while adjusting for age and sex. The effects of BMI SDS reduction on protein profiles were analyzed using linear regressions controlling for age, sex and treatment duration. The associations between changes in protein profiles and changes in continuous cardiometabolic traits were examined using linear regressions controlling for age, sex, treatment duration, baseline BMI SDS and change in BMI SDS. BMI SDS reduction was calculated as the difference between BMI SDS at baseline and BMI SDS at follow-up. Changes in protein profiles and cardiometabolic traits were calculated as the difference between the values at follow-up and those at baseline.

#### Prediction model

We performed feature selection for hepatic steatosis, defined as liver fat above 5%, using lasso regression in *glmnet* v.4.1-8 R package^53^. Through five-fold cross-validation repeated ten times, this analysis identified a three-protein panel that achieved the highest mean ROC-AUC. Furthermore, we evaluated the discriminative performance of three clinical used liver enzymes (ALT, AST and GGT), both individually and in combination with the protein panel using the same cross-validation method. To mitigate imbalanced class distribution, a down-sampling approach was applied to the majority class within each cross-validation fold. The statistical comparison of AUCs were conducted using DeLong’s test. These analyses were performed using the *caret* v.6.0.94^54^ and *pROC* v.1.18.5^55^ R packages.

#### Mediation

Mediation analyses were performed using the *mediation* 4.5.0 R package^56^. In the cross-sectional study, mediation analysis was performed to explore the mediating role of overweight/obesity on cardiometabolic traits. Bootstrapping with 1,000 iterations were employed to estimate direct, indirect and total effects across overweight/obesity-protein-trait triangles, adjusted for age and sex, at a significance level of *P* <5% FDR. The proportion of effect mediated from overweight/obesity through the protein was determined by dividing its indirect effect by the total effect.

In the intervention study, mediation analysis was performed to examine the mediation effect of protein changes on the associations between BMI SDS reduction and changes in cardiometabolic traits, adjusting for baseline age, sex, and treatment duration, at a significance level of *P* <5% FDR. Bootstrap confidence intervals were used to assess statistical significance of the mediation effects.

#### Comparison with the UK Biobank

We obtained access to UKB data under accession number 32683, which has received ethical approval from the National Health Service National Research Ethics Service (ref 11/NW/0382), with informed consent obtained from all participants. Olink Explore 3072 PEA, measuring 2923 proteins was performed as previously described^8^. We included British individuals with available Olink data, where self-report ethnicity was confirmed by genetics data (UKB field 22006). Proteins common to both The HOLBAEK Study and UKB were matched according to UniProt IDs. We compared effect sizes for associations between protein biomarkers and overweight/obesity (defined as BMI ≥25 kg/m^2^ vs. normal weight: BMI >18.5 and <25 kg/m^2^ in UKB) between the two cohorts. Prior to linear regression analysis in the UKB, each protein was rank-inverse normal transformed. Models were adjusted for age, sex, centre, fasting time, Olink batch, and time to assay.

## Data availability

All results from statistical and bioinformatics analysis are provided in Supplementary Tables 1–11. The mean levels of proteins are available in the GitHub repository at https://github.com/sarastinson/olink_holbaek/tree/main/median_data. In accordance with the General Data Protection Regulation (GDPR, https://gdpr-info.eu/), individual-level clinical and proteomics data generated in this study cannot be made publicly available to maintain patient confidentiality. However, proteomics datasets can be requested from the authors by contacting T.H. at. The obesity management protocol is available upon request from J.-C.H. at. Access to the data can be granted through the Danish Data Protection Agency and the ethics committee for the Region Zealand, Denmark, subject to obtaining proper approvals and adherence to patient information and data processing agreements. The response time for data requests is within 1 month. Accessing and processing the data requires compliance with the following conditions: (1) a data-processing agreement must be signed between the data controller and processor; (2) data may only be processed for statistical and scientific studies; (3) personal data must be deleted, anonymized and destroyed at the end of the investigation; and (4) data must not be shared with unauthorized third parties or individuals.

## Code availability

The code used for data analysis is available on GitHub: https://github.com/sarastinson/olink_holbaek

## Acknowledgments

We thank all volunteers and their parents who participated in The HOLBAEK Study. We also thank colleagues from the Hansen Group at Novo Nordisk Foundation Center for Basic Metabolic Research and The Children’s Obesity Clinic for fruitful discussions. In particular, we thank A. Frost Bjerre, B. Holløse, O. Troest, T. Larsen, A. Forman and T. Hvidtfeldt Lorentzen for technical assistance with sample preparation. We thank L. Skovborg Just and L. Ryborg for managing the GALAXY and MicrobLiver consortia. We acknowledge the funding agencies that supported this study: the Innovation Fund Denmark (grant no. 0603-00484B), the Novo Nordisk Foundation (grant no. NNF15OC0016544 to T.H.), the Novo Nordisk Foundation Challenge Program (grant no. NNF15OC0016692 to the MicrobLiver consortium (T.H. and A.K. in this study)).

S.E.S. was funded by the NNF Copenhagen Bioscience PhD Program (grant no. NNF18CC0033668). R.T. is funded by the NNF Copenhagen Bioscience PhD Program (grant no. NNF0069781). M.A.V.L. was supported by the Danish Heart Foundation (grant no. 18-R125-A8447 and 21-R149-A10071). D.B. was funded by the NNF Copenhagen Bioscience PhD Program (grant no. NNF17CC0026760). L.A.H. was supported by the Danish Cardiovascular Academy, which is funded by the Novo Nordisk Foundation (grant no. NNF20SA0067242) and the Danish Heart Foundation (grant no. PhD2023009-HF). C.E.F. was supported by the BRIDGE-Translational Excellence Program (grant no. NNF18SA0034956), Steno Diabetes Center Sjælland and the Region Zealand Health Scientific Research Foundation (grant no. R32-A1191). The European Union’s Horizon 2020 research and innovation program (grant no. 668031 to the GALAXY consortium (A.K., T.H., and M.T. in this study)). We also thank the Novo Nordisk Foundation for supporting the Novo Nordisk Foundation Center for Basic Metabolic Research (grant nos. NNF18CC0034900 and NNF23SA0084103).

## Contributions

The manuscript was drafted by S.E.S, Y.H., R.T., E.S. and T.H. S.E.S. performed the bioinformatics analysis and generated figures for the manuscript. Y.H., R.T. and E.S. contributed to bioinformatics analysis and results interpretation. J.-C.H. and T.H. designed and coordinated the clinical cohort. M.A.V.L., C.E.F., L.A.H. recruited participants in clinical cohorts, collected samples and clinical data. D.B., L.Ä. and T.I.A.S. supported statistical analysis. M.A.V.L., H.B.J., M.J., A.K., O.P., M.C. contributed to data coordination and management. J.-C.H. and T.H. developed the present project concept and protocol and supervised the project. All authors reviewed the manuscript before submission.

## Competing interests

H.B.J. is currently employed at Novo Nordisk. All other authors declare no competing interests.

**Correspondence and requests for materials** should be addressed to Jens-Christian Holm and Torben Hansen.

