## Extended Data Figure 1-3 for "Identification of modifiable plasma protein markers of cardiometabolic risk in children and adolescents with obesity"

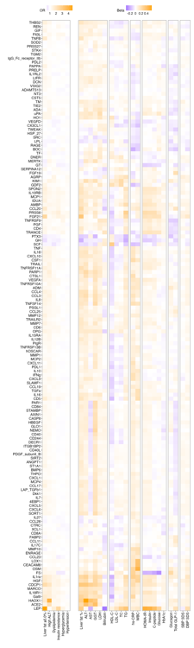

**Extended Data Fig 1.** **Associations of plasma proteins with cardiometabolic risk.** 149 proteins associated with cardiometabolic risk features and traits. Logistic regression analysis was performed adjusting for age, sex and BMI SDS. Their associations with cardiometabolic traits tested by linear regression are shown in parallel. Proteins are hierarchical clustered.

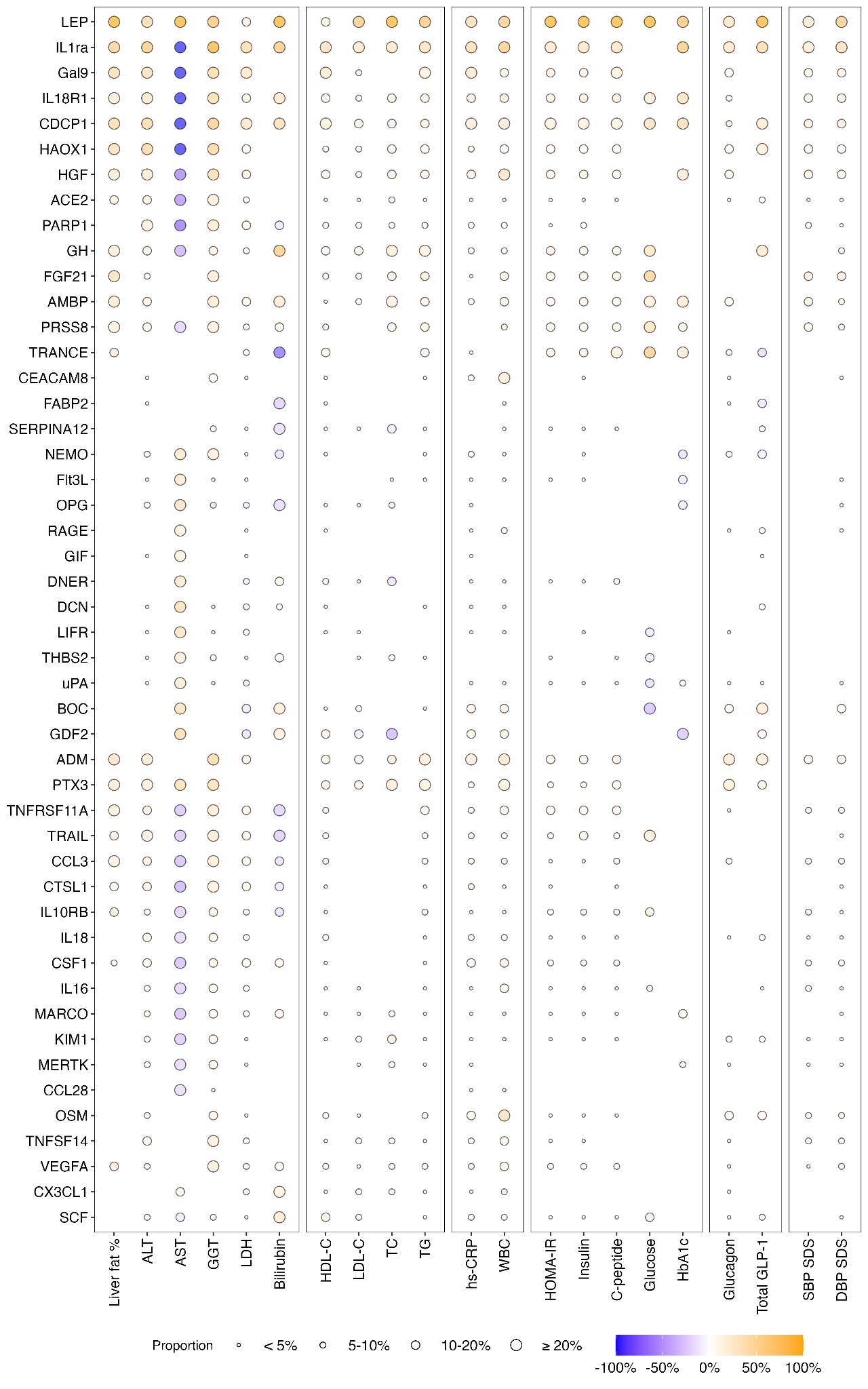

**Extended Data Fig 2. The mediation of 122 overweight / obesity-associated proteins on 21 cardiometabolic traits.** Mediation analyses were performed adjusting for age and sex. Each dot represents a significant indirect effect (*P* < 5 % FDR), with dot size indicating the mediation proportion categorized into < 5 %, 5-10 %, 10-20 %, and ≥ 20 %. Only proteins with at least one mediation proportion ≥ 20 % are displayed. Proteins are hierarchical clustered. The sample size (n) for each trait is listed in Table 1.

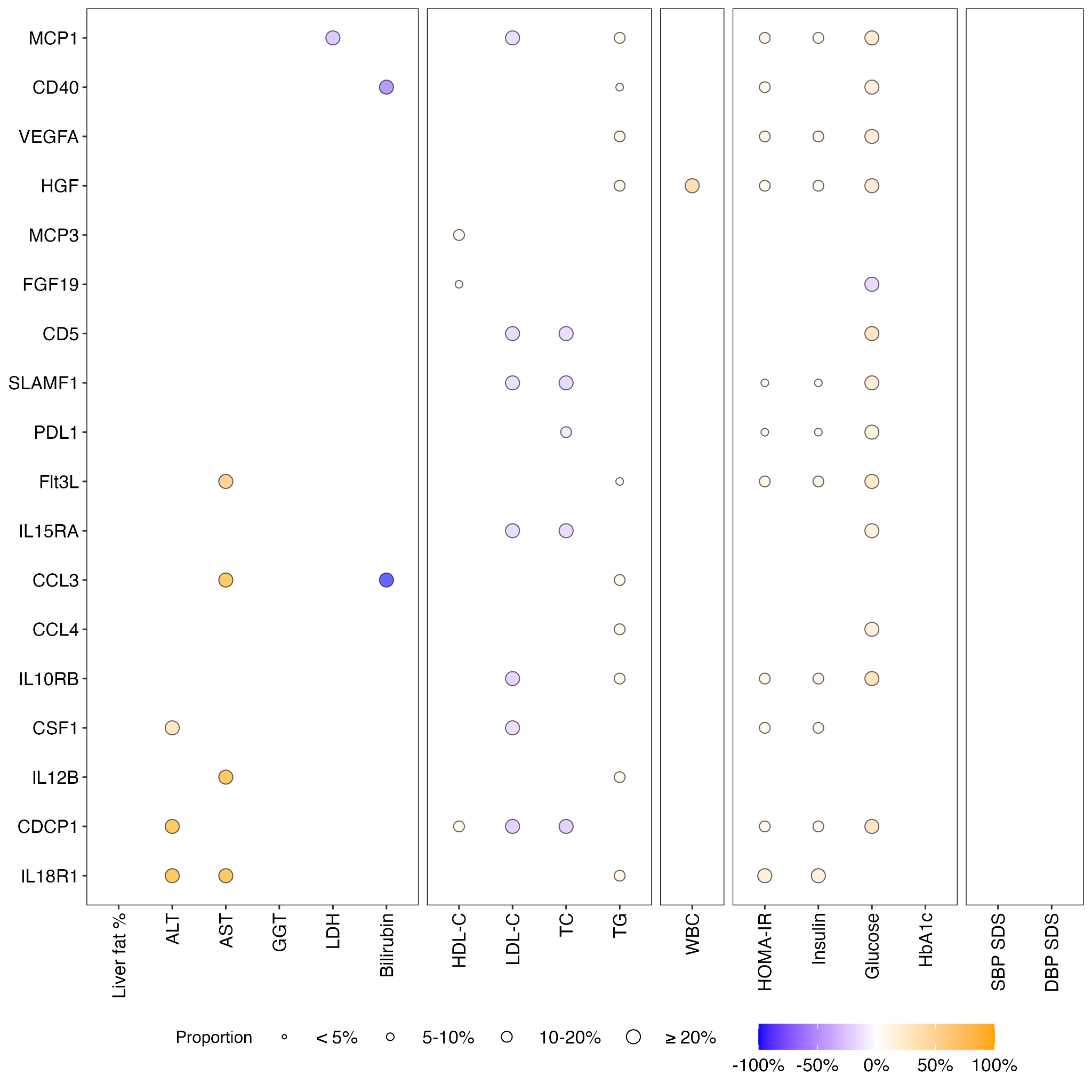

**Extended Data Fig 3. The proportion mediated by changes in 64 proteins on the association between BMI SDS and changes in 17 cardiometabolic traits.** Mediation analysis was performed adjusting for age, sex, and treatment duration. Each dot represents a significant indirect effect and proportion mediated effect (*P* > 0.05), with a dot size indicating the mediation proportion categorized into < 5 %, 5-10 %, 10-20 %, and , and ≥ 20 %. BMI SDS reduction was calculated as the difference between BMI SDS at baseline and follow-up Changes in protein profiles and cardiometabolic traits were calculated as the difference between the values at follow-up and baseline. The sample size (n) for each trait is listed in Extended Data Table 1.
